# Electroconvulsive Therapy during the COVID-19 Pandemic: Nationwide Data from Denmark

**DOI:** 10.64898/2026.02.13.26346228

**Authors:** Christian Jon Reinecke-Tellefsen, Anders Ørberg, Søren Dinesen Østergaard

## Abstract

The COVID-19 pandemic had substantial impact on healthcare systems across the globe, including psychiatric services. Use of electroconvulsive therapy (ECT), a lifesaving intervention for severe mental illness, was reported to have declined during the pandemic in several countries, but nationwide data remain scarce. Using nationwide data from the Danish National Patient Register, we examined all ECT treatments administered in Denmark from September 2019 to May 2025. Weekly treatment numbers were visualized across the three national COVID-19 lockdowns to descriptively assess changes in ECT use. A notable reduction in ECT treatments was observed in the weeks preceding and during the first lockdown (March 11 to May 18, 2020). A post-hoc estimation indicated approximately 1,366 “missed” treatments during the initial pandemic phase in 2020. When these were added to the 27,033 treatments delivered in 2020, the adjusted total approximated annual treatment volumes in 2019 and 2022, suggesting a temporary disruption rather than sustained decline. In contrast, ECT activity during the second and third lockdowns appeared largely unaffected. These findings suggest that ECT provision in Denmark was temporarily reduced during the initial phase of the pandemic but remained resilient thereafter. In the case of a future pandemic, safeguarding timely access to ECT—particularly in early phases— should be prioritized given its critical role in the treatment of severe mental illness.

## INTRODUCTION

The COVID-19 pandemic had a tremendous impact on the world and on hospital services across the globe. One of the medical specialties that was substantially affected by the pandemic was psychiatry. Specifically, the field witnessed a significant shift from in-person contacts to telemedicine for patients already in treatment (1) and substantial reductions in referrals to psychiatric treatment (2). Reports from other countries than Denmark have suggested that the, arguably, most critical treatment performed at psychiatric hospitals, namely electroconvulsive therapy (ECT), was used less during the COVID-19 pandemic (3, 4). This could potentially have detrimental consequences as ECT can be lifesaving for patients with severe manifestations of mental illness (5).

While most data on the use of ECT during the COVID-19 pandemic, with at least one notable exception from France (4), stem from case reports, surveys and small-to medium-sized cohorts (3), there are few reports with nationwide coverage - as such data are rarely available. In Denmark, however, all ECT treatments from public psychiatric hospitals (there are no private ECT providers) are reported to the Danish National Patient Register (6) and are available for research purposes. Therefore, the aim of this study was to describe ECT treatment in Denmark during the COVID-19 pandemic.

## METHODS

We extracted data on all ECT treatments in Denmark in the period from January 1, 2019 to May 18, 2025 (end of our data). ECT treatments from September 2, 2019 (well before the pandemic) to May 18, 2025 (well after the pandemic) were aggregated at the weekly level and visualized in a bar plot, while highlighting the three pandemic lockdowns that were imposed by the Danish Government, namely from March 11, 2020 to May 18, 2020, from December 17, 2020 to April 6, 2021, and (partial lockdown) from December 19, 2021 to February 1, 2022 (7). The mean number of weekly ECT treatments for the period was also visualized. We chose to take a descriptive approach to these data as a fitting statistical model was hard to identify due to the waxing and waning of the severity of the pandemic, the public measures to contain it as well as the public (patient) sentiment, all of which are likely to impact the use of ECT.

The use of register-based data for the purpose of this study was approved by Statistics Denmark and the Danish Health Data Authority. According to Danish legislation, informed consent is not required for register-based studies (waiver for this study: 1-10-72-8-26). All analyses were carried out using Sas Enterprise Guide version 8.3 via remote access to Statistics Denmark.

## RESULTS

The results are shown in Figure 1. The most pronounced observation is the gradual drop in the weekly number of ECT treatments leading up to the first COVID-19 lockdown, and the quite consistent low number of treatments throughout this lockdown. While the beginning of the second and third lockdowns also coincides with two weeks of low ECT treatment intensity, they also coincide with the Christmas and New Year holiday, which is also associated with two weeks of low ECT activity outside the COVID-19 period (only one week in 2022, where Christmas eve and New Years eve fell on Saturdays, likely limiting the impact to the week in between).

**Figure 1.**
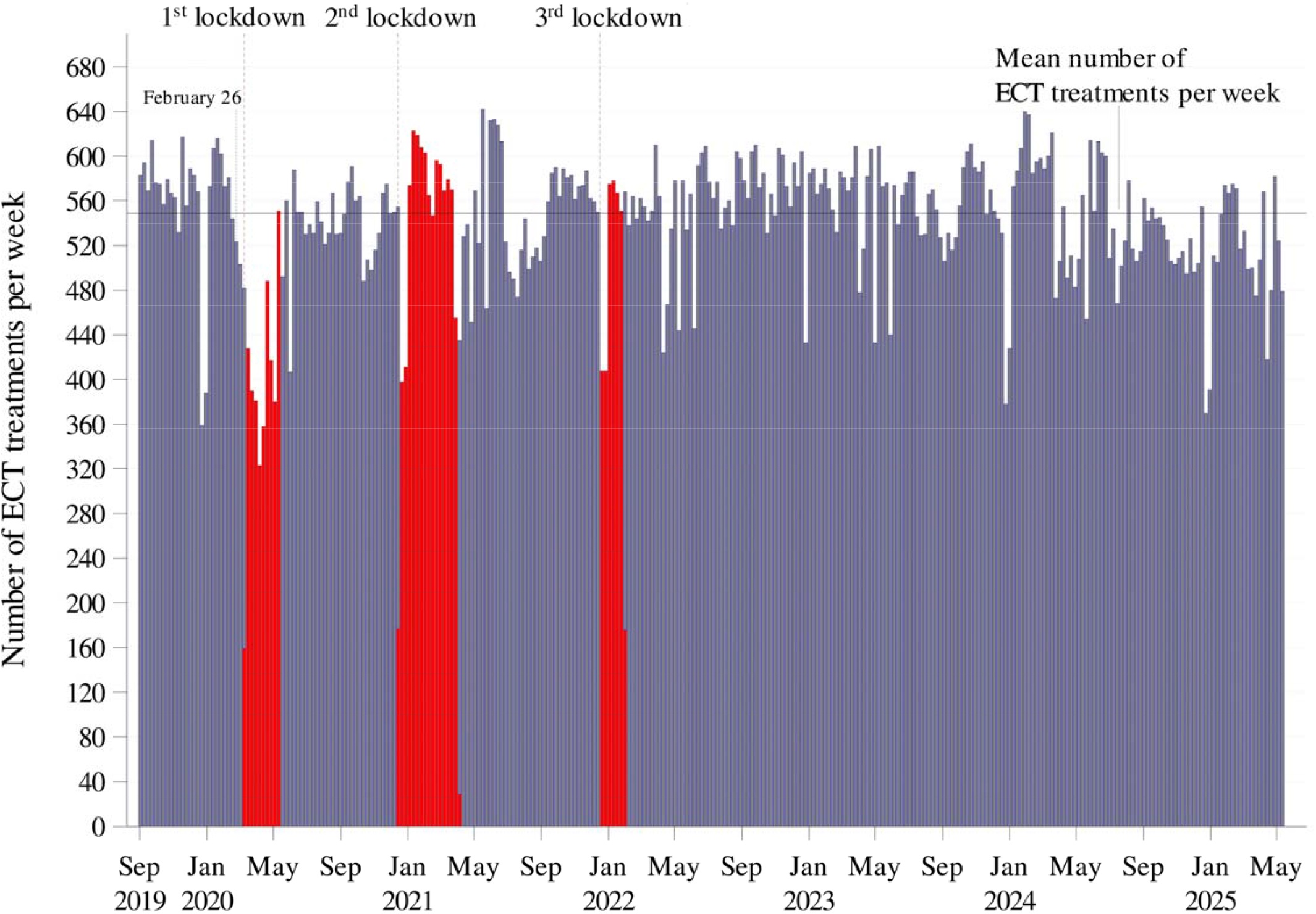
Weekly ECT treatments in Denmark from 2019 to 2025.

Based on these observations, we conducted a post-hoc analysis to estimate the quantitative impact of the first COVID-19 lockdown and the weeks of uncertainty leading up to it. Specifically, we calculated the number of ECT treatments below the mean (weekly mean: 549) in the period from February 26, 2020 (the day of the first confirmed case of COVID-19 in Denmark) to May 18, 2020 (the last day of the first lockdown) (7). This “area under the mean” amounted to 1366 “missed” treatments. When adding those to the 27,033 actual treatments in 2020, yielding 28,399 treatments, it largely corresponds to the total number of treatments in 2019 (n=27,874) and in 2022 (n=29,048), years not/little impacted by the pandemic, while accounting for a trend towards an increase in ECT use over the late 2010s and early 2020s, as also observed elsewhere (4).

## DISCUSSION

The findings of this nationwide study suggests that the use of ECT in Denmark was reduced due to the pandemic during the first COVID-19 lockdown and the time leading up to it, but also that it appeared relatively unaffected during the rest of the pandemic, including under the second and third lockdown, likely due to population habituation to- and less intense political/press rhetorics in the coverage of the pandemic.

That the COVID-19 pandemic led to decline in ECT use is in agreement with the results from most studies from other countries having investigated this aspect (3), including the nationwide study from France by Barruel et al. (4). With regard to the reasons for the reduction, the experiences from our local ECT services (Aarhus University Hospital in Denmark) from the first lockdown was that ECT capacity was not the issue, but rather that patients were less likely to seek treatment during the time. This is in keeping with results from the analysis of referrals to our psychiatric services which were reduced by 40% in the four weeks following the initiation of the first COVID-19 lockdown (2). In other countries, however, reduced ECT capacity during the pandemic seems to have played a substantial role as well (3).

The main strength of this study is the fact that it is based on longitudinal nationwide data that we consider to be highly valid, given that reporting is strictly mandatory. The longitudinal aspect proved particularly important to not mistake the drop in ECT use during the beginning of the second and third lockdown for a consequence of the lockdowns, as it appears to be explained by a drop in ECT treatments taking place every year around Christmas and New Year. The main limitation of the study is its observational nature, which prevents us from making strong causal claims, as we have no knowledge of the counterfactual (the level of ECT usage in the period in question had the COVID-19 pandemic not taken place). It could be argued that employing interrupted time-series analysis, as done by Barruel et al. in their analysis of the effect of the COVID-19 pandemic on ECT use in France (4), would have improved causal leverage. Specifically, to assess “the effect of the COVID-19 pandemic on ECT use”, Barruel et al. employed interrupted time-series analysis with time unit defined as month and the following time segmentation: “first pre-pandemic period (T1) from January 1, 2017 to February 29, 2020; a second large pandemic period (T2) from March 1, 2020 to December 31, 2022; and a third post-pandemic period (T3) from January 1, 2023 to December 31, 2023.” Judged by our data, however, an interrupted time-series approach with this predefined time segmentation would not allow for sufficiently fine-grained results to adequately understand what likely took place in Denmark with regard to ECT treatment during the COVID-19 pandemic. Another limitation is that the results from the present study may not generalize well to countries without a tax-funded public hospital system where ECT usage is free for patients, and the ECT use per capita is relatively high.

In conclusion, this study suggests that use of ECT in Denmark was reduced due to the pandemic during the first COVID-19 lockdown and the time leading up to it, but that the use appeared relatively unaffected during the rest of the pandemic, including under the second and third lockdown. In case of a future pandemic, ensuring access to ECT, particularly in its initial phase, should be given priority due to the lifesaving potential of this treatment.

## Data availability statement

The data cannot be shared due to restrictions enforced by Danish law for protecting patient privacy.

## Acknowledgements

None

## Author contribution

All authors contributed to the design of the study. SDØ procured the data. Analyses were performed and the results were visualized by AØ. The results were interpreted by all authors. CR-T and SDØ wrote the initial draft of the manuscript, which was revised by important intellectual content by AØ. All authors approved the final version of the manuscript prior to submission.

